# AI Tool Implementation to Assist with Management of Patient Portal Messages: Auto-Categorization and Routing

**DOI:** 10.64898/2026.10.19.26365557

**Authors:** Julianne D. Brooks, Maria A. Donahue, Jhansi Anjana Rayapureddy, Bradley Molyneaux, Elyse R. Park, Louisa G. Sylvia, Rebecca G. Mishuris, Andrea H. Thurler, Lidia M.V.R. Moura

## Abstract

**Background and Objective:** The volume of patient portal messages has grown substantially and artificial intelligence (AI) can support clinical staff and improve the timeliness of patient care. Our objective is to test if an AI tool for managing the electronic inbox “In Basket” reduces response times to patient portal messages in neurology.

**Methods:** We conducted an implementation study from January to May 2026 in the neurology department at two clinical sites within a regional medical system. A single AI tool was implemented to auto-categorize and auto-route all patient messages received in the electronic inbox. The tool also generates optional draft replies to messages. Primary process measures were the median weekly time (hours) from message creation to the first action and to closure. Impact was analyzed using a controlled interrupted time-series (ITS) design, comparing message-volume-matched control divisions.

**Results:** Among 24,142 messages analyzed, pre-intervention median weekly time to first action was 11.3 hours [IQR: 5.6, 15.1] (Site A) and 16.1 hours [IQR: 13.8, 19.0] (Site B), falling post-intervention to 1.0 hour [IQR: 0, 3.1] (Site A) and 0.6 hours [IQR: 0.2, 2.4] (Site B). Controlled interrupted time-series analysis showed a significant immediate differential level change in median weekly hours to first action at Site A (−92.5%, 95% CI: −97.5% to −78.0%), and a significant downward trend at Site B (−43.5%, 95% CI: −65.2% to −8.2%). Changes in time to closure were directionally favorable but not statistically significant at either site. There was low use of the generated draft replies, only 0.5% at Site A and 3% at Site B.

**Discussion:** Implementation of AI-assisted message categorization and routing was associated with a substantially reduced initial action time for patient In Basket messages in Epilepsy. The utilization of AI-generated draft-replies to messages was low. These findings support the feasibility and effectiveness of AI-augmented workflows to improve inbox management.

**TAKE HOME POINTS:**

- AI-augmented In Basket tools for categorization and routing of messages may substantially accelerate initial action on patient portal messages by directing messages to the appropriate staff.
- There was low utilization of AI generated draft-replies.
- Auto-categorization and routing alone was not associated with reducing time to closure of messages.

## INTRODUCTION

Patient-facing portal technology has fundamentally altered ambulatory medicine.^1,2^ Since the onset of the COVID-19 pandemic, clinicians have received approximately 57% more patient portal messages, a surge that has occurred without proportionate increases in staffing or reimbursement.^3^ Clinicians in the top quartile for electronic inbox volume face more than six times the odds of burnout compared with those in the lowest quartile, making inbox management one of the most potent predictors of professional exhaustion in clinical practice.^3,4^

Electronic health record (EHR) event log data confirm that ambulatory physicians spend substantial time outside scheduled clinic hours on inbox tasks, including reviewing, triaging, and responding to patient messages.^5,6^ For neurologists in particular, high-volume patient portal engagement is common, as patients with neurological diagnoses (e.g., epilepsy) use online portals at high rates and often submit messages about urgent or complex clinical concerns, including seizure activity, medication side effects, and functional decline.^7^

Existing strategies to mitigate portal-related inbox burden include redistributing message work through team-based triage models, implementing standardized communication and routing protocols, and, more recently, billing for eligible portal message time as asynchronous e-visits.^8,9^ While these approaches have achieved incremental gains in some settings, their impact has been limited by implementation complexity, workflow heterogeneity, and the need for sustained behavioral change across clinical teams.^10^ Structural interventions, such as delegation frameworks and role clarification, have shown promise in reducing raw message volume in primary care,^10^ but have not consistently accelerated response times.^11^

Emerging generative artificial intelligence (AI) and EHR-integrated AI tools can automatically and accurately categorize patient portal messages and generate context-aware draft replies, allowing existing care teams to triage and respond more efficiently to high volumes of patient inquiries without adding staff. At the same time, clinicians remain responsible for final review.^12–14^ However, AI-augmented Basket tools have not been evaluated in a neurology subspecialty setting, and no objective improvements in time-based response metrics have been demonstrated in other settings.

We evaluated the impact of implementing an AI-powered tool for message auto-categorization, auto-routing, and draft-reply generation in the EHR In Basket,^12^ focusing on time-based response outcomes in two clinical neurology sites within a regional system.

## METHODS

### Standard Protocol Approvals, Registrations, and Patient Consents

This report follows the Enhancing the Quality and Transparency Of Health Research (EQUATOR) guidelines for quality improvement studies (SQUIRE 2.0; **eTable 1**).^15^ This study received approval from the Mass General Brigham (MGB) Institutional Review Board. The requirement for informed consent was waived because the data analyzed in this study were collected as part of routine clinical care, de-identified, abstracted retrospectively, and aggregated for analysis.

### Study Design

We conducted an implementation study from January to May 2026, with 12 weeks of pre-implementation and seven weeks of post-implementation at two neurology clinical sites, or Brigham and Women’s Hospital (Site A) and Massachusetts General Hospital (Site B) within the regional medical system, Mass General Brigham (MGB). The AI tool for auto-categorization, auto-routing, and draft-reply generation for patient messages was implemented across the neurology epilepsy division. We included messages for individuals who had prior encounters in the neurology department. Only messages written in English were included. Patient messages are routed to one of three groups (i.e., nursing, front desk, or prescription) or directly to clinicians. We included all patient-initiated messages directed to clinicians in the neurology department at both Sites A and B and then selected messages for clinicians in the epilepsy division and two other divisions as controls. We selected control divisions based on pre-intervention weekly message volume and ensured that nursing staffing was similar across groups. We included messages that had a completed action or were closed at the time of data collection, with a one-week lag.

We accessed patient portal message records from the MGB Enterprise Data Warehouse. We included messages from January 11, 2026, to May 1, 2026, allowing for 12 weeks of baseline data and 7 weeks of follow-up post-intervention. For Site B, we truncated the data at four weeks post-intervention due to staffing changes that affected the number of nurses responding to pool messages.

### Pre-Intervention Workflow

A diagram of pre- and post-intervention workflows for Site A and B is shown in **eFigure 1**. Before the intervention, Sites A and B had different workflows and types of staff responding to patient portal messages. At Site A, messages were either screened by patient service coordinators (PSCs) or nurses. At Site A, the epilepsy (target) division had one full-time equivalent (FTE) epilepsy nurse covering the message pool and the control divisions had either one FTE nurse (neuroimmunology) or PSCs (comprehensive pool) covering the message pools. PSCs handled non-clinical messages and forwarded clinical messages directly to physicians, whereas nurses handled messages or escalated to physicians. At Site B, the message pools were covered by nurses for all divisions, with 1.5 FTE for epilepsy, 2 for movement disorders and 2 for neuroimmunology. Prior to the intervention at Site B, nurses screened messages and forwarded non-clinical messages to front desk. Clinical messages were handled by designated nursing staff or escalated to physicians. The steps to message closure may vary by site and by division based on whether they have nursing support to handle messages (**eFigure 1**). In general, the steps for handling messages were to 1) screen the message, 2) forward to the appropriate recipient, 3) complete actions to handle the request 4) reply to the patient and 6) mark the message as done. Auto-categorization and routing can address screening and forwarding of messages (Steps 1 & 2) whereas draft-replies can help with replying to patients (Step 5).

**Figure 1.**
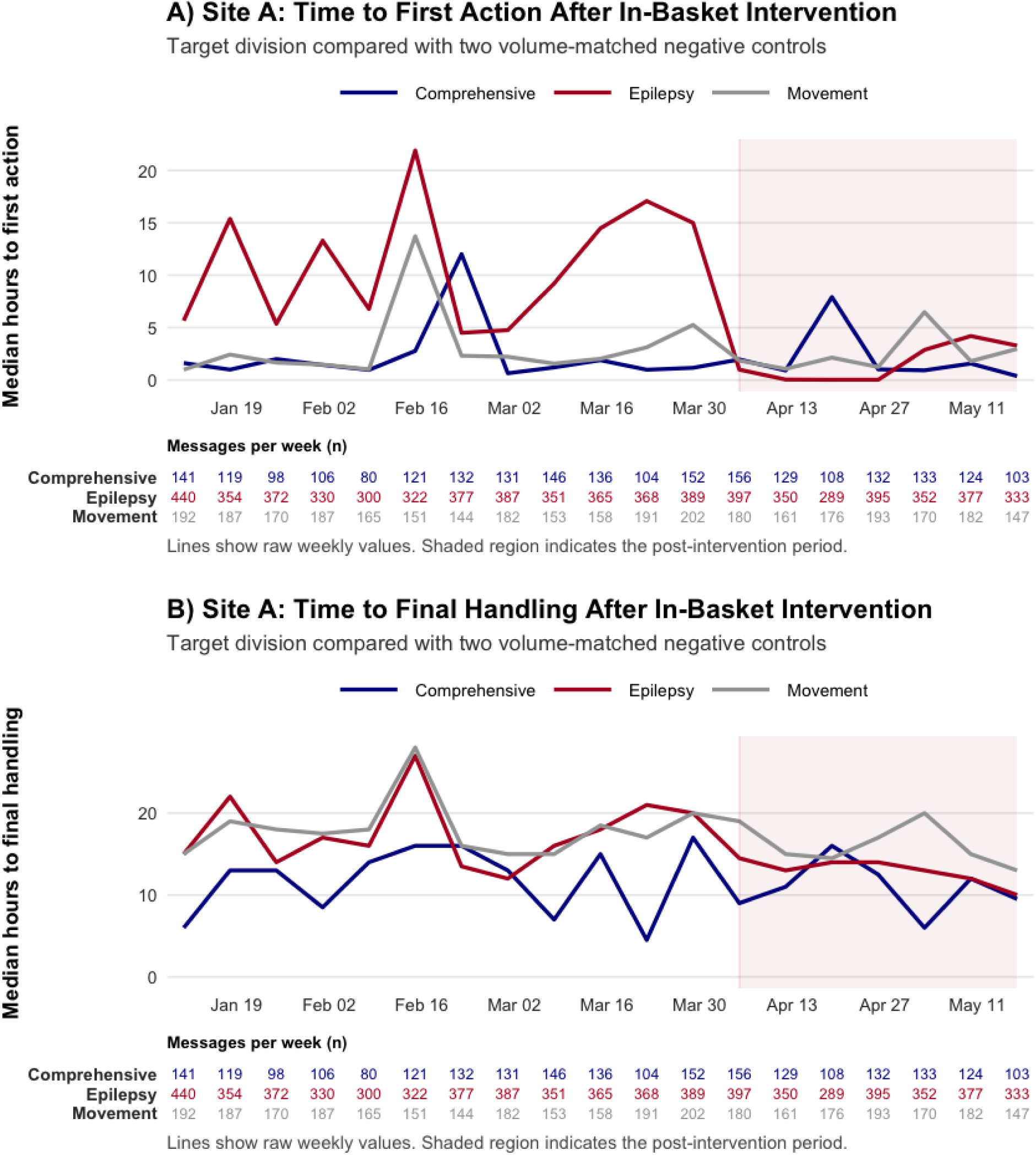
Site A Time to First Action and Final Handling After In Basket Intervention. **Legend:** Site A Weekly A) Time to First Action and B) Time to Final Handling of In Basket Messages Pre and Post-Implementation of Auto-Categorization and Draft Replies.

### Post In Basket Tools Intervention Workflow

An AI tool was implemented in the Epic electronic inbox “In Basket” to assist with patient message management. On April 12th, 2026, we implemented Epic’s In Basket PMAR Categorization and Augmented Response Technology (ART), which is a set of AI tools which automatically characterized and routed messages and generated draft responses to patient messages. All tools for categorization, routing and draft replies were developed and tested by Epic. Model performance documentation can be found in Epic Galaxy In Basket PMAR Categorization Model Brief^16^ and further details are available in EPIC Galaxy under the articles “Get Medical Advice Setup and Support Guide” subheadings: Enable Predictive Models for In Basket Categorization and Route In Basket Messages More Efficiently.^17^ For auto-categorization, messages were sorted by a predictive machine-learning model that uses cognitive computing to review the message text and apply a relevant category. The messages were first categorized into clinical or non-clinical messages and then assigned a detailed category. The categories for clinical messages were: medication, lab, procedure, referral, advice request, paperwork, COVID-19, results or authorization (**eTable 2**). The categories for non-clinical messages were scheduling, billing, medical info, tech support, gratitude and acknowledgement. After being assigned a category, messages were routed to the appropriate pool using a rule-based system. Clinical messages were routed to clinical pools (staffed by nurses or clinicians) and non-clinical messages were routed to front desk pools. For draft replies, the tool automatically generated a response to the patient message based on the message text and information from the patient record, such as current prescriptions. When responding to a message, clinical staff reviewed and revised a draft response or started a blank reply. The draft-reply feature had very low uptake (less than 5% of message responses). During the course of the study, we gathered qualitative feedback from end-users and asked staff to report any patient safety issues.

### Study Measures

The unit of analysis was patient-initiated clinical messages. Messages created before April 12, 2026, were categorized as “pre-intervention,” and messages created after that date as “post-intervention.” Primary outcomes were the time from message creation to first action (hours) and to closure (hours), aggregated weekly as median values. We used median weekly values, as the time to action was right skewed, with a small number of data points having a long time to first action/closure. The first action indicates how the message was first acted on in the audit trail. The options for first action are no further action, called patient, forwarded message, message reply, message reply from visit navigator, message merge, simple thank you, new, read, pending, hold and done. Time to closure tracks the time for the message to be marked as done. Secondary measures included the percent of messages handled within 24, 48, and 72 hours.

### Statistical Analysis

We analyzed the impact of the intervention using a controlled interrupted time-series (ITS) analysis, comparing the epilepsy division to two message volume–matched divisions within the neurology department as negative controls. Message volume for the target division compared to matched controls is shown in **eFigure 2**. We used linear regression with weights adjusted for message volume and included indicators for pre- and post-intervention, the sequential study week, and the sequential week post-intervention for the target and control divisions. Interrupted time-series analysis accounts for pre-intervention trends and allows us to compare to control divisions. The analysis was conducted using R Version 2026.01.0+392.

**Figure 2.**
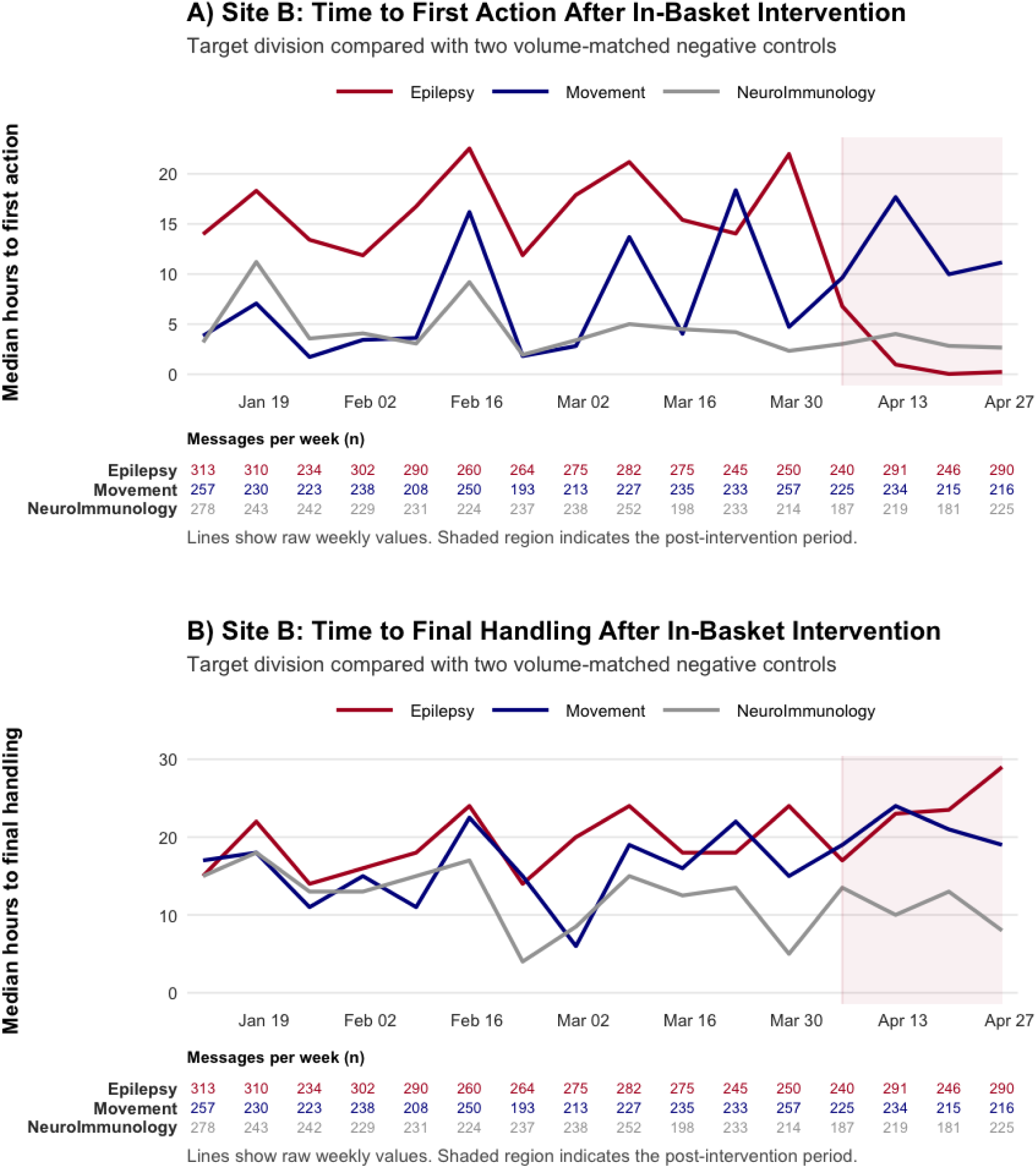
Site B Time to First Action and Final Handling After In Basket Intervention. **Legend:** Site B Weekly A) Time to First Action and B) Time to Final Handling of In Basket Messages Pre and Post-Implementation of Auto-Categorization and Draft Replies

### AI Use

The authors used Anthropic Claude Opus 4.6 in the code creation and editing for figures. Authors JDB and LVMRM are responsible for all analysis and reviewed every line of code, can explain what each section does and can interpret the output. All data represented in the tables and figures were verified through data quality checks completed by the authors.

## RESULTS

We analyzed 24,142 patient messages. Tables 1 shows the message volume pre- and post-intervention message volume for the epilepsy, movement (control), and comprehensive neurology (control) divisions for Site A, as well as the epilepsy, neuroimmunology (control), and movement (control) divisions for Site B. Epilepsy was the highest-volume division at both sites.

Pre-intervention, median weekly time to first action was 11.3 hours (Site A) and 16.1 hours (Site B); median time to closure was 16.5 hours (Site A) and 18.0 hours (Site B). Post-intervention, median time to first action (i.e., either a response or forwarding to another team member) fell to 1.0 hours (Site A; Figure 1) and 0.6 hours (Site B; Figure 2). Post-intervention time to closure was 13.0 hours (Site A) and 23.5 hours (Site B). Draft reply at Site A from April to May was 0.5%, and in Site B, 3.37%.

Controlled ITS (Table 2) showed a significant immediate differential level change in median weekly hours to first action at Site A (−92.5%, 95% CI: −97.5% to −78.0%, p<0.001). At Site B, the immediate differential level change was not statistically significant (−63.1%, 95% CI: −91.3% to 56.3%, p=0.176), but there was a significant downward post-intervention trend (−43.5% per week, 95% CI: −65.2% to −8.2%, p=0.021). Changes in time to closure were directionally negative, but did not reach statistical significance at either site.

**Table 1.** Patient Portal Message Volume and Response Times.

|  | Pre-Intervention | Post-Intervention | Pre-Intervention | Post-Intervention | Pre-Intervention | Post-Intervention |
| --- | --- | --- | --- | --- | --- | --- |
| Division | Messages<br>(median/wk) | Messages<br>(median/wk) | Median<br>Time to<br>First<br>Action<br>(hrs) | Median<br>Time to<br>First<br>Action<br>(hrs) | Median<br>Time to<br>Closure<br>(hrs) | Median<br>Time to<br>Closure<br>(hrs) |
| **Site A (Brigham and Women's Hospital)** |  |  |  |  |  |  |
| **Epilepsy\*** | **4,355 (367)** | **2,439** | **11.3** | **1.0** | **16.5** | **13.0** |
| Movement Disorders** | 2,082 (176) | 1,209 | 2.1 | 1.9 | 17.8 | 15.0 |
| Comprehensive Neurology** | 1,466 (126) | 885 | 1.3 | 1.0 | 13.0 | 11.0 |
| **Site B (Massachusetts General Hospital)** |  |  |  |  |  |  |
| **Epilepsy\*** | **3,300 (275)** | **1,067** | **16.1** | **0.6** | **18.0** | **23.2** |
| Neuroimmunology** | 2,819 (235) | 812 | 3.8 | 2.9 | 13.2 | 11.5 |
| Movement Disorders** | 2,764 (232) | 890 | 3.9 | 10.6 | 15.5 | 20.0 |

**Legend:** hrs = hours; wk = week; N/A = not separately reported post-intervention. Site B data truncated at 4 weeks post-intervention due to staffing changes. Response time data for control divisions are not reported individually (used only for ITS modeling). \*Intervention, \*\*Controls

**Table 2.** Controlled Interrupted Time-Series Analysis: Effect of AI Tools on Median Weekly Time to First Action and Time to Closure.

|  |  |  |  |  |
| --- | --- | --- | --- | --- |
| Site / Outcome | Analysis Component | Differential % Change (95% CI) | p-value | Interpretation |
| *Site A (Brigham and Women's Hospital)* |  |  |  |  |
| Time to First Action | Immediate level change | −92.5% (−97.5% to −78.0%) | < 0.001 | Significant immediate reduction |
| Time to Closure | Immediate level change | Directionally negative | NS | Not statistically significant |
| *Site B (Massachusetts General Hospital)* |  |  |  |  |
| Time to First Action | Immediate level change | −63.1% (−91.3% to 56.3%) | 0.176 | Not statistically significant |
| Time to First Action | Post-intervention trend (per week) | −43.5% (−65.2% to −8.2%) | 0.021 | Significant downward weekly trend |
| Time to Closure | Immediate level change | Directionally negative | NS | Not statistically significant |
| 573<br>574<br>575<br>**Legend:** CI = confidence interval; NS = not significant. ITS = interrupted time-series. All estimates represent differential changes (intervention vs. control). Site B data is limited to 4 weeks post-intervention. R Version 2026.01.0+392 used for all analyses. |  |  |  |  |

Over the course of the study we received positive feedback on the message categorization and routing from epilepsy nurses handling messages. No patient safety events were reported during the intervention period.

## DISCUSSION

In this implementation study, the activation of AI-powered message auto-categorization and routing tools in the EHR In Basket was associated with dramatic reductions in time to first action for patient portal messages. Changes in time to closure were directionally favorable at Site A, but did not reach statistical significance at either site. There was low uptake of AI-generated draft replies to respond to patient messages.

The epilepsy division at both sites had longer time to first action compared to controls, which may be due to a larger volume of messages at baseline. At Site B, although the target and control divisions had similar nursing coverage, with 1.5 FTE nurse for epilepsy and 2 FTE nurses for the matched control divisions (movement disorders and neuroimmunology), epilepsy had a higher volume of messages at baseline per nurse. Auto-categorization helped with screening messages, a task normally completed by nursing staff, as shown by the reduction in time to first action. Despite having different operational process flows, both sites saw a reduction in time to first action and no significant change in time to closure. At Site A, there was an immediate change and at Site B there downward weekly trend in time to first action. One explanation for this difference is that the post-intervention follow-up at Site B was shortened to four weeks due to staffing changes that altered the configuration of the nursing personnel responding to the message pools. This shortened observation window reduced our ability to observe a sustained level shift. At both sites, the auto-categorization was automatically applied to all messages, so adoption of the tool should be similar across sites. However, we acknowledge there could be difference in how quickly the staff adapted to the new tool. Site B had a slight reduction in time to first action in the first week post-intervention followed by a much larger reduction in subsequent weeks, suggesting there was about a 1-week period for the intervention to take full effect.

Our study had low use of AI-generated draft replies, which were optional for nurses and providers to use when responding to patient messages. A study of patient satisfaction with AI-generated responses found higher satisfaction with AI responses compared to clinician responses to patient medical questions.^18^ Participants surveyed on their perspective of AI-drafted portal messages saw AI-tools as mutually beneficial as long as AI drafts were reviewed and approved by the clinician.^19^ Although draft replies could improve patient satisfaction and help with turn-around time, there are also many other steps in the process of handling message which may limit the efficacy. A previous study conducted in primary care and gastroenterology settings showed no objective reduction in message response time after the implementation of AI-generated draft replies alone.^20^ However, the study demonstrated improvements in clinician-reported burden and burnout with draft-reply utilization rates of 20%.^20^ Our study introduces auto-categorization and routing as an additional intervention component. We observed a reduction in time to first action, suggesting that message categorization may be the primary driver of faster initial responses. Draft-reply generation may support clinician well-being, reduce cognitive load and increase patient satisfaction, while categorization may be the critical feature for improving triaging and inbox management.

A prior study demonstrated that workflow restructuring and explicit role clarification can reduce the volume of patient portal messages in an internal medicine clinic.^10^ Their approach was to develop best standard practices for message handling and routing. They implemented a routing guide to clarify physician and staff roles and restructured staffing to have one staff member dedicated to address portal messages one morning per week. This intervention reduced message volume between team members and decreased redundant routing. Similarly, our AI-augmented workflow approach targeted the routing process of incoming messages. Both approaches could reduce the burden of incoming messages and the time required to manage each one.

To our knowledge, this is the first study to evaluate AI-assisted In Basket tools in a Neurology subspecialty setting.^21^ Prior work has been concentrated in primary care, internal medicine, gastroenterology, and emergency care, with high patient volume and different patient complexity. The work in other sub-specialties is applicable to neurology because many of the same common issues apply with categorizing urgent and non-urgent messages. For example, in primary care, patients may send urgent messages about chest pain or questions about inhalers whereas in neurology and epilepsy, many portal messages pertain to seizure activity, antiseizure medication management, and side-effect monitoring.^7,20,21^ Primary care may receive messages on a wide variety of medical topics, whereas neurology may receive more complex care management messages. This study strengthens the generalizability of AI-augmented workflows in ambulatory subspecialty medicine by demonstrating significant improvements in initial action time in this complex clinical population.^22^

In our setting, the most plausible mechanism underlying the dramatic reduction in time to first action is that automated categorization permits near-instantaneous classification and routing of incoming portal messages into clinically meaningful categories, bypassing the delays inherent in manual sorting and triage. By the time a care team member opens the message queue, each message has already been labeled as clinical or non-clinical and assigned a subcategory and routed to the correct pool. Auto-categorization and routing removes a cognitively demanding triage step that previously required reading and interpreting each message (before routing or responding). The result is a shorter time from message arrival to initial action, not necessarily full resolution, but the critical first contact that signals receipt and begins the care pathway. The use of automated tools also may have financial implications as less time is spent screening and forwarding messages, which may require less staff dedicated to inbox management.

Auto-categorization and routing reduced time to initial action, but did not have a significant effect of time to closure. Our study had low adoption of AI draft replies, which may have helped with resolution of messages. However, it is likely that time to message resolution is more dependent on downstream factors. Message closure typically requires downstream clinical decision-making processes (e.g., prescription changes, care coordination, writing seizure action plans or direct physician review) that remain human-dependent and cannot be substantially accelerated by categorization or draft generation alone.^23,24^ In the current version, the draft reply does not take into consideration the full context of the patient history and therefore the responses are limited. It is also possible that the seven-week post-intervention observation window was insufficient to capture full effects on closure time, particularly if clinicians and staff required time to adapt their workflows to the new tools.

The volume of patient electronic messages increased during the COVID-19 pandemic and never returned to pre-COVID-19 levels.^25^ Electronic message volume has been shown to be related to burnout and increased after-hours clinical work for physicians.^26,27^ One factor that contributes to burnout is having insufficient time to see patients and keep up with inbox management and documentation responsibilities. Reducing the cognitive burden of initial message triage may support better work-life balance for clinical staff, particularly nurses and advanced practice providers who shoulder a disproportionate share of inbox management responsibilities.^28–30^ If auto-categorization can shorten the reactive, repetitive dimensions of inbox work, clinicians may experience reduced after-hours burden and greater perceived control over their workload (factors known to moderate burnout risk).^4,23,31^

Importantly, no incidents of clinically consequential misrouting or other major workflow errors were reported by the nursing team during the study period. Front-desk staff noted receiving patient “thank-you” messages that, under the prior workflow, may have been directed to the nurse, advanced practice provider, or physician who had previously assisted the patient. This routing was intentional: because these messages generally required no clinical action, the AI system directed them to front-desk staff, and they were therefore not considered misrouted. Nevertheless, the absence of reported major errors does not eliminate the possibility of less apparent AI-generated errors, including misclassification, inappropriate routing, or inaccurate or hallucinated draft content, even within a human-in-the-loop workflow. A pre-intervention safety and escalation process remained in place after implementation and was reinforced through weekly team review. Future implementations should additionally maintain a structured error log to prospectively capture suspected AI hallucinations, routing errors, inappropriate draft responses, and near misses, thereby permitting estimation of their frequency and supporting continuous monitoring and workflow refinement.

The reduction in time to first action from approximately 11-16 hours to <1 hour represents a meaningful improvement in the time to screen messages and is a first step to improving inbox management. Auto-categorization and routing alone did not improve response times in our study. A future direction of this work is to reduce time to closure for urgent messages through categorization and triaging. In an epilepsy context, where patients may be reporting new seizures, medication intolerance, or worsening neurological symptoms, faster clinician response is essential and could lead to better outcomes for patients.

Key strengths of this study include the use of a controlled interrupted time-series design, which is more rigorous than an uncontrolled before-and-after comparison and allows for estimation of differential effects relative to concurrent trends in matched controls. The study was conducted in a real-world clinical environment across two academic hospital sites, encompassing a large total message volume and a clinically complex subspecialty population. The use of objective, time-stamped EHR data reduces the risk of reporting bias.

## Limitations

### Study Design

The study was conducted in the epilepsy division of the neurology department at two sites, which limits generalizability to other specialties, institutions, EHR platforms, or staffing models. The post-intervention follow-up period was brief (four to seven weeks), and long-term effects on response time, workflow adaptation, and clinical outcomes remain unknown. The 12-week pre-intervention baseline, while providing reasonable statistical stability, may capture secular trends in message response times that could confound the estimated intervention effect.

The study did not include pre- and post-intervention qualitative surveys assessing staff perceptions of workload, usability, cognitive burden, burnout, or satisfaction. Such measures could have provided complementary insight into the user experience and perceived effects of the intervention. However, survey-based outcomes are inherently more subjective than the objective, time-stamped EHR measures used in this study and may be affected by selection, recall, and recency biases, as well as differential response rates. Although prior studies of AI-assisted In Basket tools have relied extensively on clinician-reported outcomes, our controlled interrupted time-series analysis directly measured changes in actual workflow performance across a large volume of messages. We therefore consider the study findings robust, while recognizing that future evaluations combining objective workflow outcomes with systematically collected pre- and post-intervention subjective input would provide a more comprehensive assessment of both operational impact and staff experience. Additional information would be beneficial to collect in future studies such as patient satisfaction and experience, financial implications, and additional safety outcomes.

### AI and Technology Limitations

In this pilot, AI tools were applied only to English-language messages, potentially introducing differential effects for patients who communicate in other languages. However, related work on multilingual medical chatbots and AI-based translation systems demonstrates that this technology can be extended to handle multilingual communication, offering a path to address language-related access barriers in healthcare.^32,33^ Draft replies are based on the text of the message sent by the patient and some information from the patient records such as prescription and recent results. The current version does not use the full patient clinical history, which is a limitation for complex messages.

### Clinical Workflow Limitations

Auto-categorization and routing can assist with aspects of inbox manage, but the current workflow still relies on staff to act on and respond to patient messages. Data from Site B were shortened to four weeks due to mid-study staffing changes, reducing statistical power at that site. There was low use of AI-generated draft replies in our study, which limited our ability to test the impact of this intervention on time to first action and message closure.

## CONCLUSION

AI-powered auto-categorization and routing tools implemented in the Epic EHR In Basket were associated with faster initial action on patient portal messages in an epilepsy division, reducing the median time to first action from more than 11 hours to less than one hour. This study demonstrates AI tools can assist with improving inbox management in the Neurology setting. These findings support adoption of AI workflow tools in ambulatory neurology and provide a framework for future investigations measuring response time outcomes.

## Data Availability

We have full access to all the data used for this study. Anonymized data not published within this article will be made available by request from any qualified investigator.

## ACKNOWLEDGEMENTS

This study was funded by the Baker Family Initiative in Patient Experience and Outcomes. J, Brooks curated the data, conducted the formal analyses, prepared the original manuscript draft and contributed to manuscript review and editing. M, Donahue prepared the original manuscript draft and contributed to manuscript review and editing. J, Rayapureddy assisted with data curation. B, Molyneaux coordinated project implementation and administration and contributed to manuscript review and editing. E, Park, L, Sylvia and R, Mishuris contributed to manuscript review and editing. A, Thurler coordinated project implementation and administration and contributed to manuscript review and editing. L, Moura provided supervision, secured funding and resources, conceived and designed the study and methodology, validated the analytical approach and findings, and critically reviewed and revised the manuscript. All authors participated in the interpretation of the results, provided critical feedback on the manuscript, and approved the final version for publication.

## Ethics Statement

This report follows the guidelines for the Revised Standards for QI Reporting Excellence. This study received approval from the Mass General Brigham (MGB) Institutional Review Board. The requirement for informed consent was waived because the data analyzed in this study were collected as part of routine clinical care, de-identified, abstracted retrospectively, and aggregated for analysis.

