## Supplemental Material for "AI Tool Implementation to Assist with Management of Patient Portal Messages: Auto-Categorization and Routing"

***Artificial Intelligence Tools to Improve Response Time to Patient Portal Messages: Auto-Categorization and Draft Replies in a Neurology Department***

### FIGURES

#### Figure S1. Flowchart for Pre and Post-Intervention In-Basket Workflow

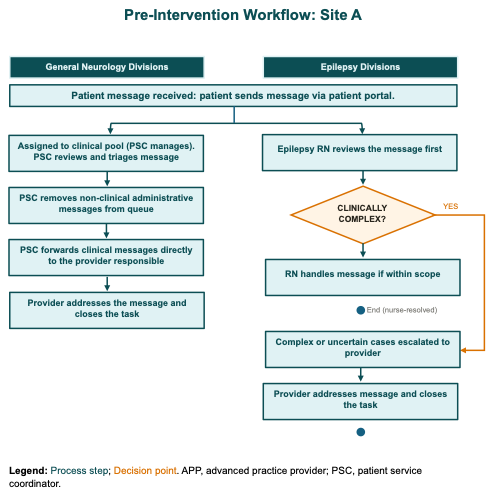

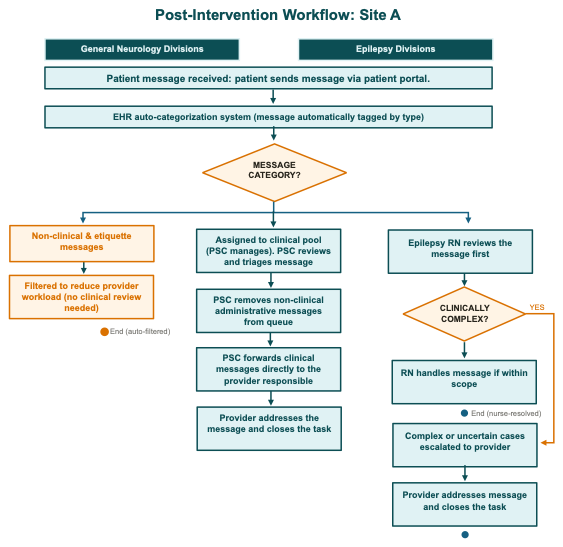

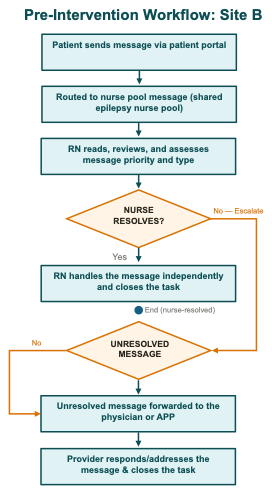

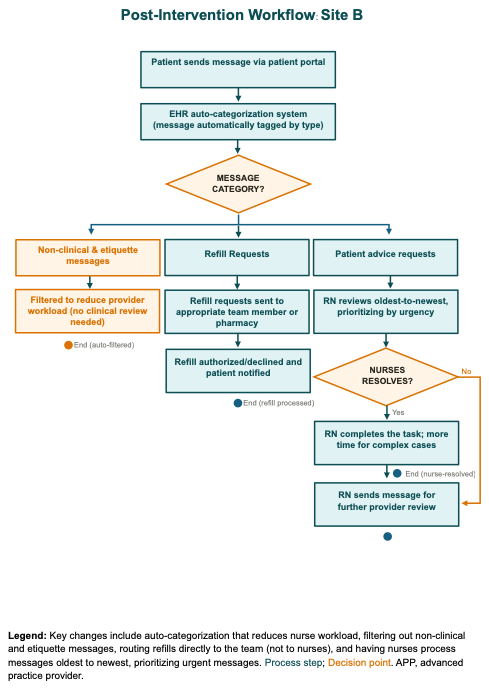

**Legend:** Key Features include consistent nurse pool routing across all divisions. 1.5 nurses cover epilepsy messages. Coverage Groups A & B ensure continuity during staffing gaps. Nurses resolve when possible; providers handle the rest. Key changes include auto-categorization that reduces nurse workload, filtering out non-clinical and etiquette messages, routing refills directly to the team (not to nurses), and having nurses process messages from oldest to newest, prioritizing urgent messages. Process step; Decision point. APP, advanced practice provider; PSC, patient services coordinator.

#### Figure S2. Message Volume for the Epilepsy Division and Two Volume Matched Control Divisions

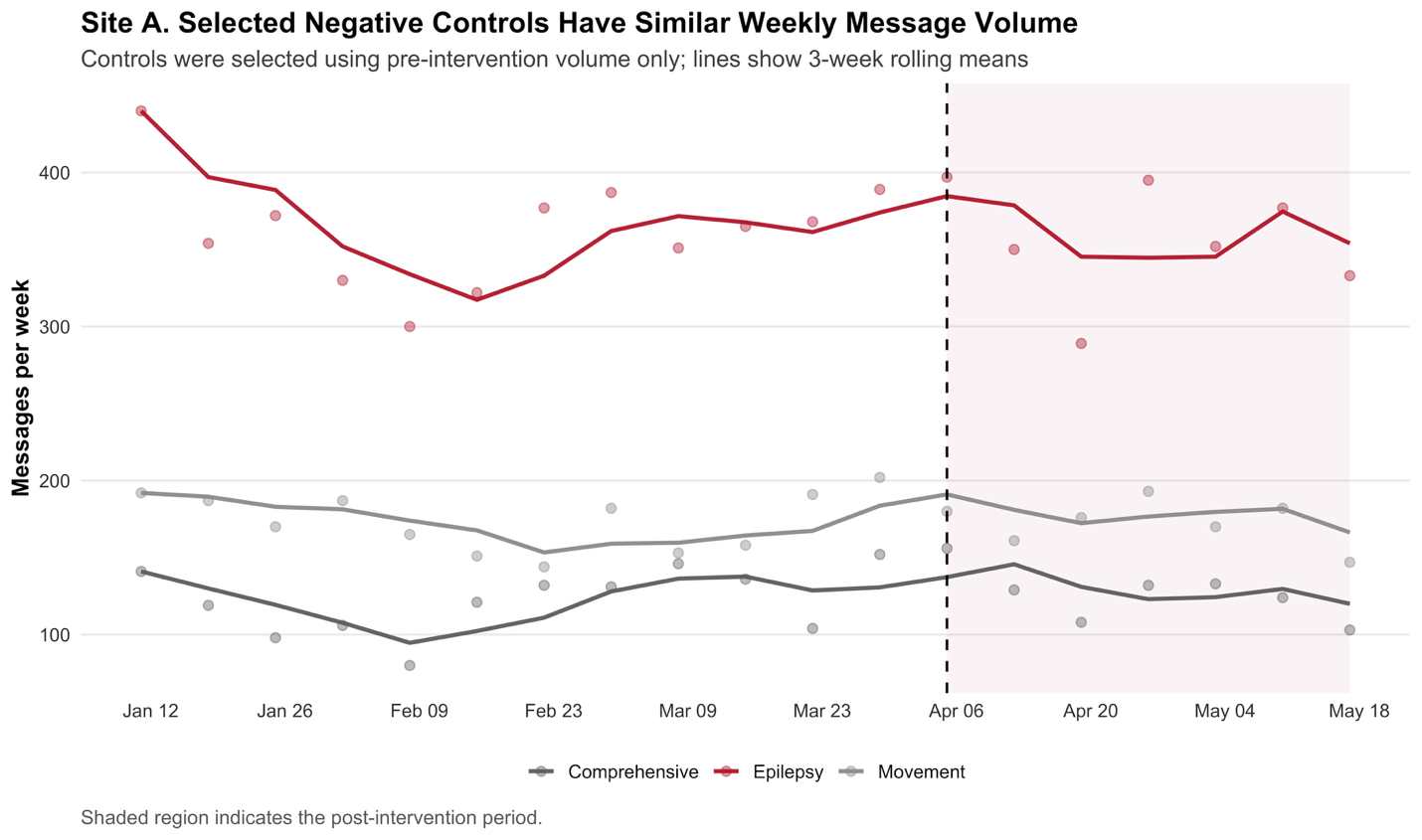

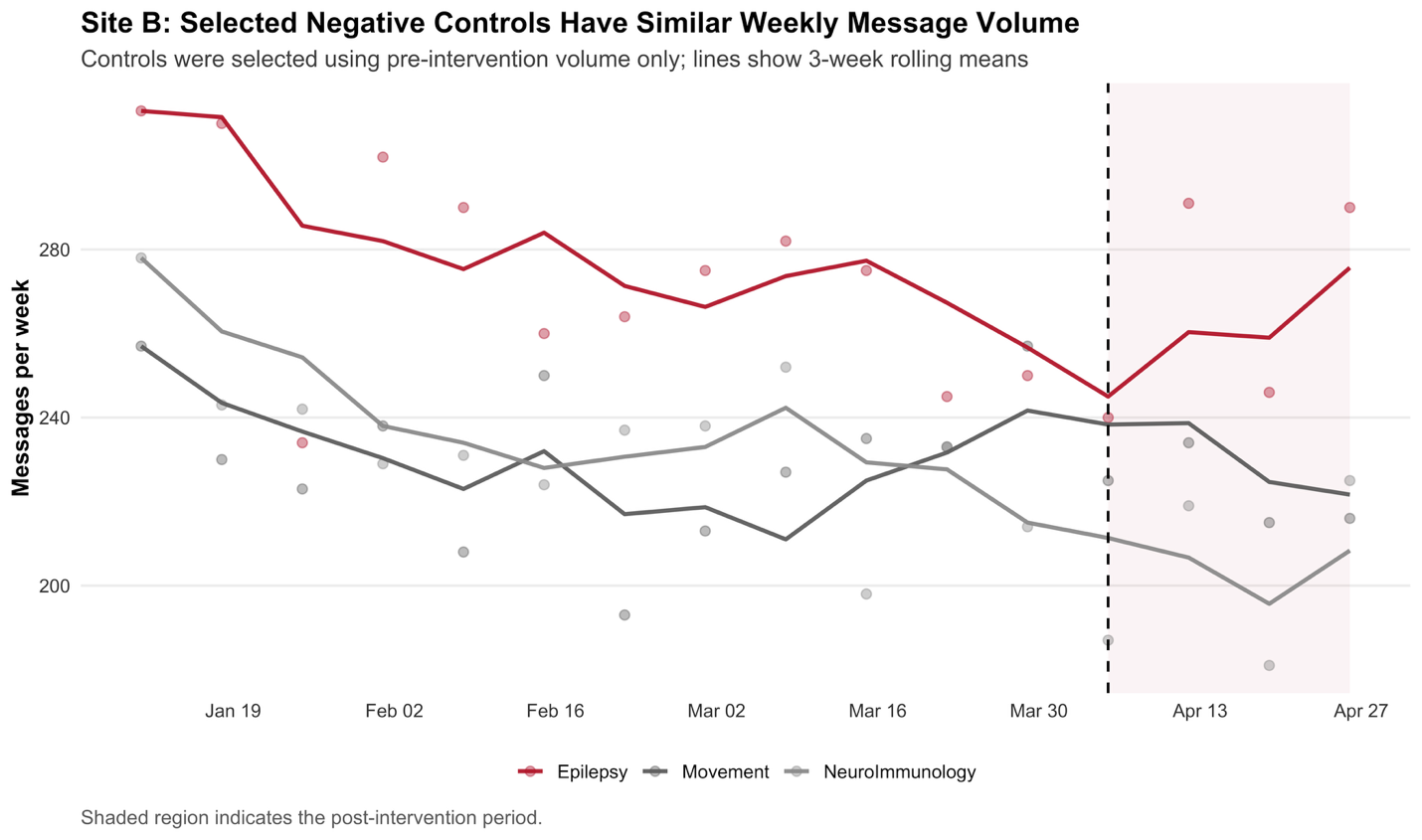

### TABLES

#### Table S1. SQUIRE Checklist

| **Section** | **Item** | **Recommendation** | **Location** |
| --- | --- | --- | --- |
| Title/Abstract | 1 | Indicate study design; summarize methods and results | Abstract |
| Introduction | 2 | Explain scientific background and rationale | Introduction |
| Objectives | 3 | State prespecified hypotheses | Introduction |
| Methods | 4 | Describe study design, setting, and participants | Methods |
| Variables | 7 | Define outcomes, exposures, and covariates | Methods |
| Bias | 9 | Describe efforts to address potential sources of bias | Methods |
| Study size | 10 | Explain how study size was determined | Methods |
| Statistical methods | 12 | Describe statistical methods, models, and sensitivity analyses | Methods |
| Participants | 13 | Report numbers at each stage of the study | Results; Table 1 |
| Descriptive data | 14 | Provide participant characteristics | Results; Table 2 |
| Outcome data | 15 | Report outcome summaries | Results; Figures 1–2 |
| Main results | 16 | Give effect estimates and precision | Results; Table 3 |
| Discussion | 18 | Summarize key results and limitations | Discussion |
| Generalizability | 21 | Discuss external validity | Discussion |
| Funding | 22 | Describe funding sources and role of funders | Funding statement |

#### Table S2. Category Definitions

| **Category 1** | **Category 2** | **Definition** |
| --- | --- | --- |
| Clinical | Clinical - General | Requires clinical follow-up, like a message requesting a refill or asking for medical advice. |
|  | Clinical Advice | Requires a clinician to respond with medical advice. This includes worsening symptoms (even if the patient does not explicitly ask for advice) and general questions about a new problem. |
|  | Authorization | Messages about prior authorization |
|  | Lab - Question | Messages about labs |
|  | Lab - Request | Request for a lab order |
|  | Medication - Question | Messages about current or new medications |
|  | Medication - Request | Request for a new (or refill-expired) medications |
|  | Referral | Messages about referrals. This label is relatively rare-- questions about location, time and contact should be labeled as Scheduling. |
|  | Referral Request | A request for referral, including questions like "should I see …". Questions about location, time and contact should be labeled as Scheduling. |
|  | Result | Messages about results findings or values |
|  | Result Request | Request to view a result |
|  | Paperwork | Message requesting clinician fill in some paperwork, such as a work excuse, school excuse, or FMLA documentation. |
|  | COVID-19 | Messages about COVID-19 |
| Non-Clinical | Nonclinical – General | Requires administrative follow-up, such as messages about billing or scheduling. |
|  | FYI | Does not require follow-up. |
|  | Scheduling | A question about an upcoming appointment like location, time, or contact info (including labs, procedures, and referrals) or a request to cancel/reschedule an appointment. |
|  | Billing | A question about a hospital bill or insurance coverage of some service requested or rendered |
